# Optimal Respiratory Syncytial Virus intervention programmes using Nirsevimab in England and Wales

**DOI:** 10.1101/2022.08.24.22279152

**Authors:** David Hodgson, Mihaly Koltai, Fabienne Krauer, Stefan Flasche, Mark Jit, Katherine E. Atkins

## Abstract

**Introduction:** Respiratory Syncytial Virus (RSV) is a major cause of acute lower respiratory tract infections (ALRI) in infants. There are no licensed vaccines and only one monoclonal antibody available to protect infants from disease. A new and potentially longer-lasting monoclonal antibody, Nirsevimab, showed promising results in phase IIb/III trials. We evaluate the cost-effectiveness of Nirsevimab intervention programmes in England and Wales

**Methods:** We used a dynamic model for RSV transmission, calibrated to data from England and Wales. We considered a suite of potential Nirsevimab programmes, including administration to all neonates (year-round); only neonates born during the RSV season (seasonal); or neonates born during the RSV season plus infants less than six months old before the start of the RSV season (seasonal + catch-up).

**Results:** If administered seasonally to all infants at birth, we found that Nirsevimab would have to be priced less than £63 per dose for at least 50% certainty that it could cost-effectively replace the current Palivizumab programme, using an ICER threshold of £20,000/QALY. An extended seasonal programme which includes a pre-season catch-up becomes the optimal strategy below a £34/dose purchasing price for at least 50% certainty. At a purchasing price per dose of £5-32, the annual implementation costs of a seasonal programme could be as high as £2 million before a switch to a year-round strategy would be optimal.

**Discussion:** Nirsevimab has the potential to be cost-effective in England and Wales not only for use in high-risk infants.

## INTRODUCTION

Respiratory Syncytial Virus (RSV) is a common seasonal virus which causes acute lower respiratory tract infections (ALRI), including bronchiolitis and pneumonia. In children younger than five years, RSV is the most common cause of ALRI, a leading cause of morbidity and mortality globally in this age group.[1]. During 2015–2016, estimates for the global annual burden of RSV-related ALRI in children under five years of age are 25.4–44.6 million cases, 2.9– 4.6 million hospitalisations and 101,400 deaths.[2,3]. There are limited pharmaceutical options to protect infants against RSV infection. Only one prophylactic is licensed, Palivizumab; however, it is expensive, and its protection is short-lived, requiring up to five monthly doses to protect against disease throughout a season.[4]. Consequently, Palivizumab is administered only to extremely vulnerable, premature infants in high-income countries.[5]These factors prompted the WHO in 2015 to highlight RSV vaccine development as a high-priority global health goal.[6] In response, there are currently 22 RSV vaccines in clinical trials, varying in the target group (e.g. infants, pregnant women, elderly) and vaccine type (e.g. Inactivated, recombinant, live-attenuated). [7]

One of the most promising prophylactics in development is Nirsevimab, a single-dose long-acting monoclonal antibody, being developed by AstraZeneca and Sanofi.[8] The results of the MELODY trial, a phase II/III randomised control trial assessing the safety and tolerability of Nirsevimab, suggest a higher efficacy against hospitalisation (62% vs. 55%) and a longer duration of protection (∼150 days vs 30 days) than Palivizumab.[4,9,10]. The prospect of a more effective prophylactic against RSV, which is expected to be cheaper than a single dose of Palivizimab,[11] could broaden access to RSV prevention from only high-risk neonates to healthy infants who also have a large burden of RSV disease.[12] The Joint Committee on Vaccination and Immunisation (JCVI) in the UK has emphasised that modelling should be used to explore the impact of expanding the eligibility criteria for RSV immunisation with this new monoclonal antibody.[13] However, to maximise health benefits under ever-tightening national healthcare budgets, the scope of an augmented RSV intervention programme needs to be carefully evaluated using cost-effectiveness analyses (CEA).

In this study, we evaluate the purchasing price per dose (PPPD) for large-scale Nirsevimab immunisation strategies to be cost-effective in England and Wales.

## METHODS

### RSV transmission model structure

We used a previously published model of RSV transmission in England and Wales to evaluate the epidemiological impact, of Nirsevimab programmes.[14] We used an SEIR transmission model stratified into 25 age groups (monthly up to 11 months of age, and then 1, 2, 3, 4, 5–9, 10– 14, 15–24, 25–34, 35–44, 45–54, 55–64, 65–74, 75+ years).

We assumed that infected individuals can be symptomatic or asymptomatic, and that asymptomatic infections are less infectious compared to symptomatic infections.[15] Additionally, we assumed that infants are born with waning maternally-protective antibodies and that natural immunity gradually builds up over 3 RSV infections, reducing the risk/severity of a further infection each time an individual becomes infected. Finally, we assume that the risk of getting infected depends on contact between people in different age groups, informed using empirical data from England and Wales.[16,17] We captured seasonal effects in the transmission of RSV by multiplying the per-contact transmission rate with a seasonal forcing term. The seasonality of RSV in England and Wales has been largely unpredictable between 2020 and at the time of writing due to restrictions on social mixing due to the COVID-19 pandemic.[18,19] We make the simplifying assumption in this model that once Nirsevimab is licensed the observed seasonality prior to 2020 will return.

### Economic model

Our economic model translated the age-stratified RSV incidence from the transmission model into five RSV-associated: symptomatic infection, GP consultations, hospital admissions, hospital bed days, and deaths. The health benefit was measured in terms of the quality-adjusted life-years (QALYs) gained by each intervention programme, calculated using UK-specific estimates for QALY loss for symptomatic cases, hospitalised cases and deaths.[20] The cost of each programme was calculated in 2018 GBP, from the perspective of the National Health Service. Primary care consultations and hospital bed days were used to estimate the cost of the burden of RSV, and the implementation cost for each programme was the sum of each vaccine’s purchasing cost and administration cost.[21–23] All costs and effects were discounted at a rate of 3.5% over a 10-year time horizon.[24]

Full details of the transmission and economic models can be found in a previous publication.[14]

### Intervention programmes

#### Status quo

We assumed that Palivizumab has an efficacy of 74.3% (95% Credible Interval (CrI) 56.6–85.7), provides protection throughout the RSV season, and is currently administered to 90% of Very high-risk infants at birth between October to February inclusive, as per current Palivizumab guidelines for England and Wales.[25]

#### Long-acting monoclonal antibody (Nirsevimab)

We tracked the number of infants protected by long-acting monoclonal antibodies which remain protected after birth for an average of 150 days (with an exponential loss of protection) with an efficacy against RSV disease of 74.5% (95% CrI 49.6–87.1%) as observed for Nirsevimab.[9] We evaluated five programmes that administer a single dose of long-acting monoclonal antibodies (i) to those infants who are currently eligible for Palivizumab at birth (Very high-risk), (ii) to all infants born from October to February at birth (Seasonal), iii) to all children at birth year-round (Year-round) (iv) to all infants born October to February at birth in addition to all infants less than 7 months of age in October (Seasonal with catch-up), and (v) to all infants born October to February at birth in addition to infants at either 8, 12, or 16 weeks of age (to coincide with the existing National Immunisation Programme (NIP) in England) at the closest time to the start of the RSV season (Seasonal with NIP-integrated catch-up) (**Table 1**). We assume that either of these programmes would replace the existing Palivizumab programme and that they all achieve the same coverage as the Palivizumab programme.

**Table 1.** Summary of the intervention programmes parameters.

| Programme | Eligible population | Window | Coverage | Annual # doses |
| --- | --- | --- | --- | --- |
| Very high-risk | Very-high-risk infants | Oct-Feb | 90% | 1,458 |
| Seasonal | All infants at birth | Oct-Feb | 90% | 213,481 |
| Year-round | All infants at birth | All year | 90% | 525,676 |
| Seasonal with catch-up | All infants at birth | Oct-Feb | 90% | 506,306 |
|  | Infants aged 1-5 months | Oct | 90% |  |
| Seasonal with NIP integrated catch-up | All infants at birth | Sep-Feb | 90% | 514,254 |
|  | 4 months | July | 90% |  |
|  | 4 months | Aug | 90% |  |
|  | 4 months | Sep | 90% |  |
|  | 2-4 months | Oct | 90% |  |

**Table 2.**
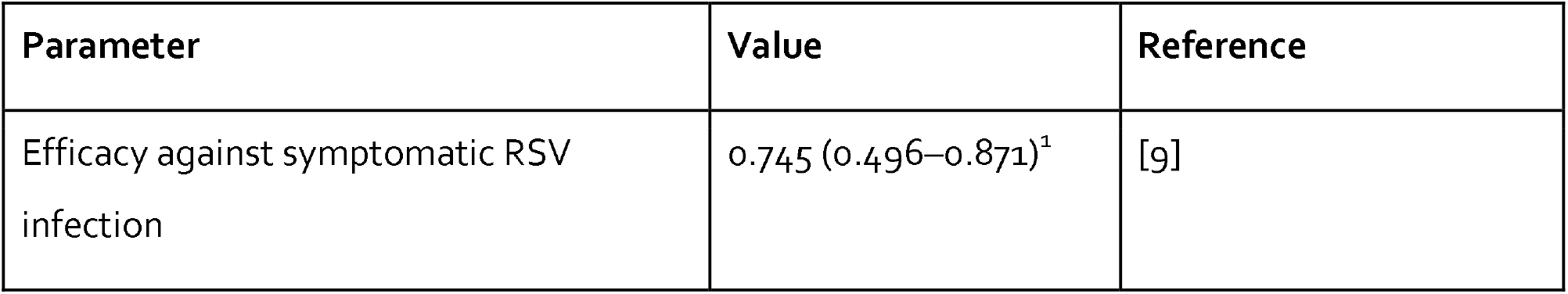
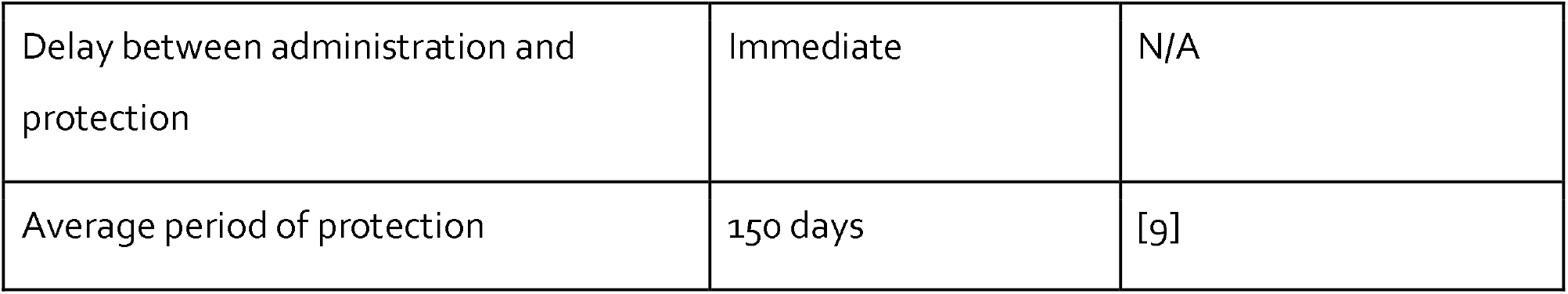
Summary of the immunological parameters associated with Nirsevimab. ^1^ Fitted distribution: Beta(12.45, 6.94)

| Parameter | Value | Reference |
| --- | --- | --- |
| Efficacy against symptomatic RSV infection | 0.745 (0.496–0.871)<sup>1</sup> | [9] |
| Delay between administration and protection | Immediate | N/A |
| Average period of protection | 150 days | [9] |

### Determining the optimal programme

We calculated the incremental cost-effectiveness ratio (ICER) of the current Palivizumab programme in England and Wales and the five Nirsevimab programmes.[25] As the total cost of each programme depends on the purchasing price per dose (PPPD), we determined which intervention programmes are dominant (i.e. both less costly and more impactful than any other strategy) or dominated, separately for a fixed PPPD. We consider PPPD values from £1– £4,600 in £1 intervals. Ignoring dominated programmes, the optimal programme is defined as the most impactful (i.e. has the greatest QALY gain) which is cost-effective (below the willingness-to-pay threshold of £20,000/QALY) compared to the next most impactful non-dominated comparator. This analysis was repeated for 1,000 Monte Carlo samples from i) the fitted posterior distributions of the dynamic transmission model, ii) samples from the credible intervals for the efficacy values, and iii) samples from the credible intervals for parameters in the economic models; including QoL and costs.

### Sensitivity analysis

We performed a sensitivity analysis on parameters to capture uncertainties associated with i) the average duration of protection of Nirsevimab (increasing from 150 days to 250 days and 360 days), ii) the coverage dropping from 90% to 70%, iii) the average age of first administration from birth to 2 months, and iv) a drop in the willingness-to-pay threshold from £20,000/QALY to £15,000/QALY (**Table S1**).

### Annual costs of seasonal administration

For highly seasonal diseases like RSV in the UK, seasonal administration is more efficient, but can often be administratively burdensome. Using our cost-effectiveness framework, we quantified the maximum annual cost that one should be willing to pay to ensure that the seasonal programmes remain cost-effective at £20,000/QALY, compared to their year-round counterpart. This maximum annual cost provides an upper limit on the sum of both the actual incremental economic cost of implementing a seasonal programme and the willingness-to-pay price for any logistical difficulties in implementing a seasonal programme. That is, if the maximum cost is below the sum of the actual cost of implementing a seasonal programme compared to a year-round programme and the cost associated with logistical issues, then switching to a year-round strategy is optimal.

## RESULTS

### Impact of programmes on healthcare outcomes

The year-round and the two seasonal catch-up intervention programmes (Year-round, Seasonal with catch-up and Seasonal with NIP-integrated catch-up) had a similar impact on preventing RSV-related healthcare outcomes (**Figure 1A**). Using hospital cases as an example; Year-round, Seasonal with catch-up and Seasonal with NIP-integrated catch-up programmes were able to prevent 6452 (95% CrI 5076–7566), 6839 (95% CrI 5353–8039), 6693 (95% CrI 5248–7861) hospitalised RSV cases across all ages, respectively; resulting a reduction of 26.7% (95% CrI 20.9–31.2), 28.6% (95% CrI 22.4–33.6), and 27.9% (95% CrI 21.8–32.7) of hospital cases for those aged 1 year and under. The Seasonal programme is the most efficient (cases averted per dose), however, preventing 16.1 (95% CrI 12.7–19.0) and 424 (95% CrI 333–496) hospital cases and symptomatic infections respectively for every 1,000 children immunised (**Figure 1B**). Comparatively, the Year-round and catch-up programmes are less efficient, preventing 10.4 (95% CrI 8.2–12.2), 11.5 (95% CrI 8.9–13.5) and 11.1 (95% CrI 8.7–13.0) of hospital cases per 1,000 doses, respectively. Most of the impact was attributable to protection in infants under 1 year of age across all programmes and all healthcare outcomes except symptomatic disease. Herd protection due to vaccination was negligible in preventing healthcare outcomes across all programmes (**Figure 1C**).

**Figure 1.**
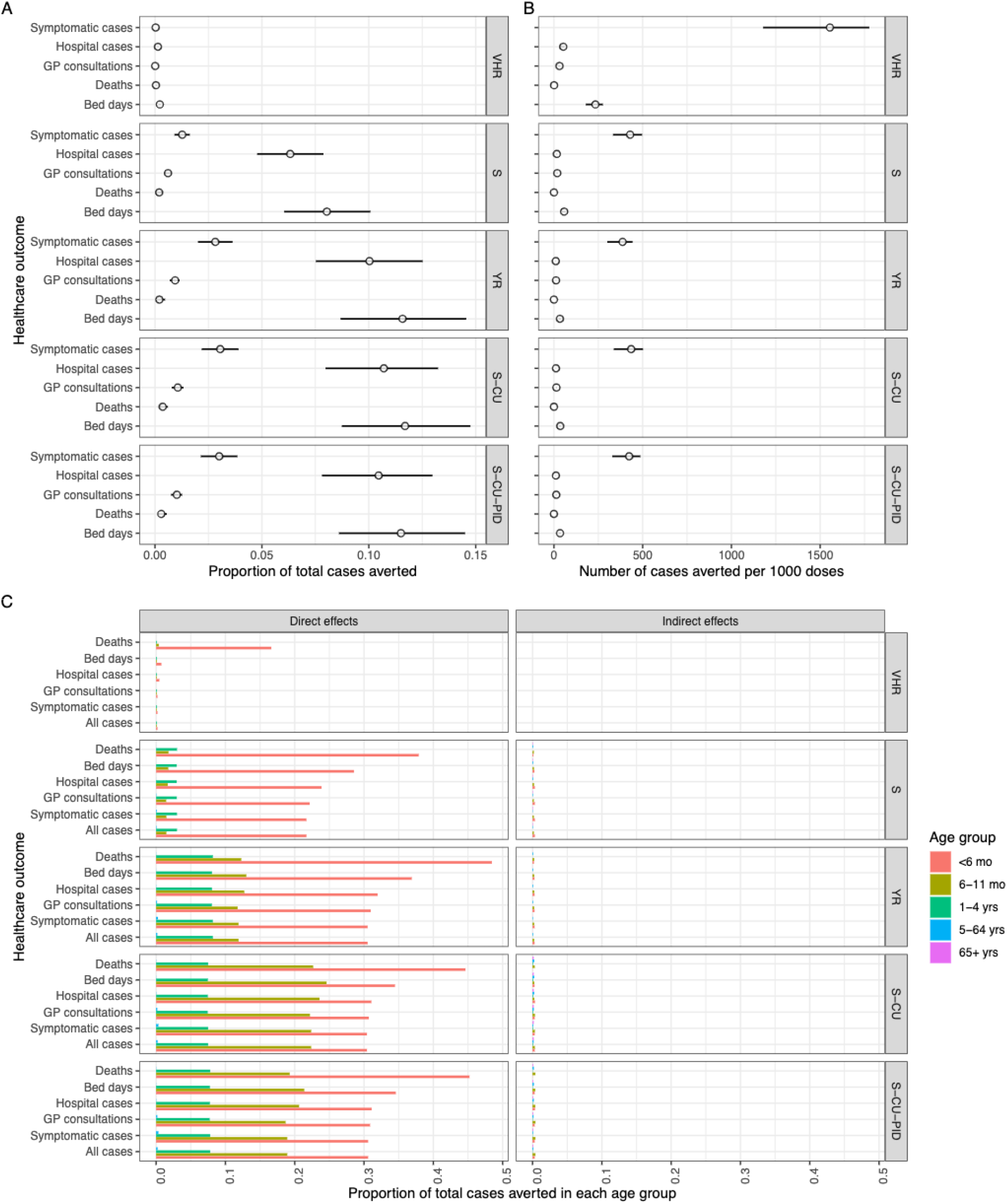
Impact of intervention programmes. The impact of the five intervention programmes for Nirsevimab. a) Proportion of health care outcomes averted (symptomatic infection, hospital admission, death, GP consultations, and bed days) across all ages for each of the five programmes. Point markers represent the mean and the line ranges represent the 95% CrI of the posterior distribution. b) Number of cases of health care outcomes averted per 1000 doses given for each programme c) Proportion of each health care outcome averted attributable to the direct effects (left panel) and indirect effects (right panel) across all ages for each of the five programmes (VHR: Very high-risk, S: seasonal, YR: Year-round, S-CU: Seasonal with catch-up, S-CU-NIP: Seasonal with NIP-integrated catch-up).

### The optimal intervention programme

The PPPD determines the total cost of each programme and, thus, which intervention programmes are dominant and dominated. Both the Seasonal with NIP-integrated catch-up and the Year-round programmes are always dominated, within the considered range of PPPD (**Figure S1**), if no additional delivery costs of the seasonal programme were assumed. The Very high-risk programme is dominated when the PPPD is less than £31, while the Palivizumab programme is dominated when PPPD is less than or equal to approximately £4,300, (**Figure S1**).

Which Nirsevimab programme is optimal (cost-effective with the largest impact) depends on the PPPD (**Figure 2a and b**). If the PPPD is less than or equal to £32 then the Seasonal with catch-up programme is optimal; between £33-63 the Seasonal programme is optimal; and over £63, the Very high-risk programme is optimal. Consequently, a PPPD larger than £63 means a vaccination programme with Nirsevimab is unlikely to be cost-effective unless only restricted to infants at very high risk of disease. However, continuing the existing Palivizumab programme but with Nirsevimab as a prophylactic agent, (i.e. the Very high-risk programme) is optimal between £63–4400 PPPD, above which the existing Palivizumab programme remains the optimal programme.

**Figure 2.**
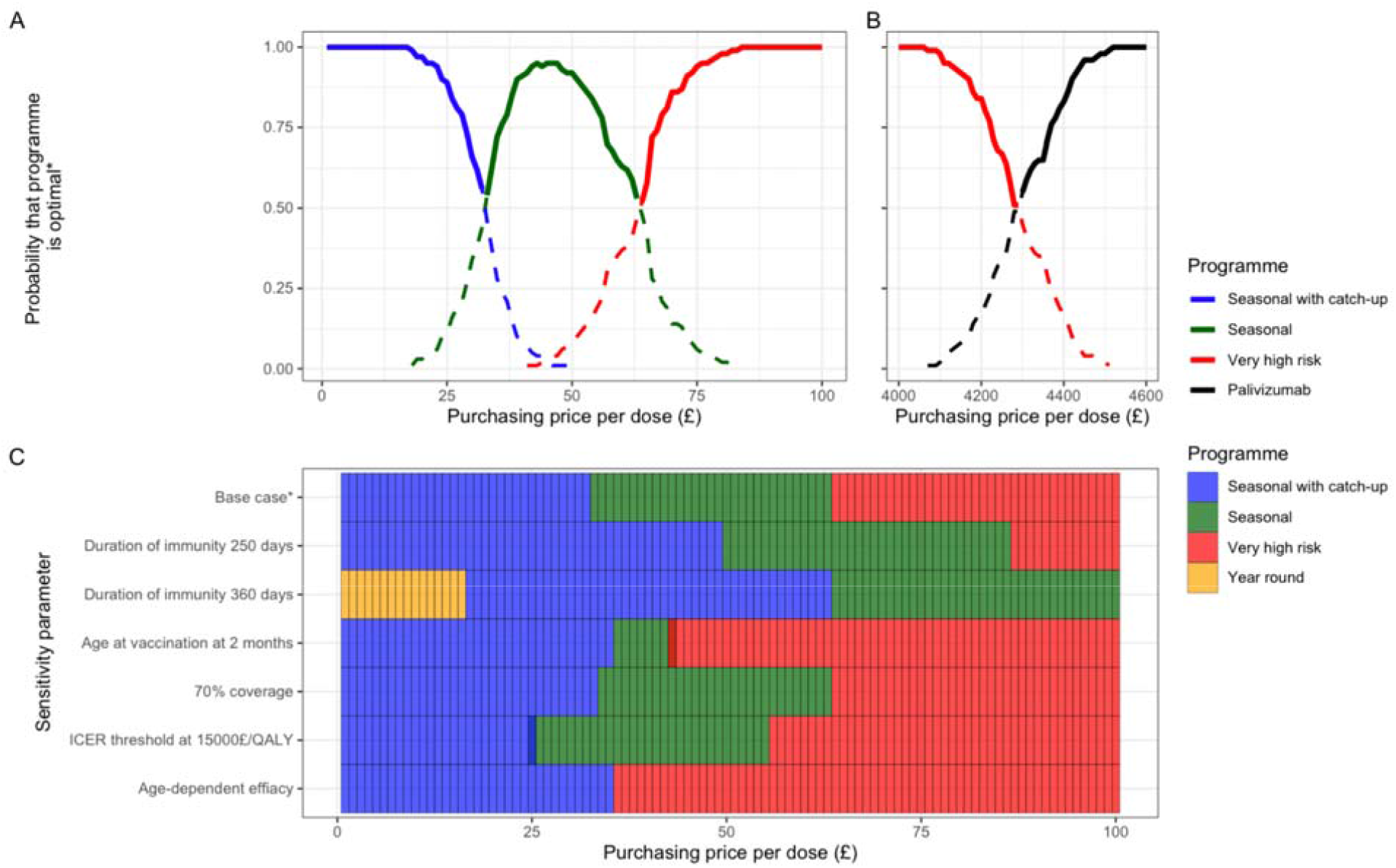
A/B Optimal intervention programmes given purchasing price per dose. The probability that an intervention programme of the base case is optimal (i.e. the cost-effective programme with the largest QALY reduction) for PPPD between £0-100 (A) and between £4,000-4,600 (B). C. The most frequently occurring optimal interval programme for PPPD between £0–100 and sensitivity parameters. The overlapping colours represent instances when either of two programmes is optimal, i.e. have an equal number of samples.

The sensitivity analysis changes which intervention programme is optimal given a PPPD. The duration of the protection granted by Nirsevimab has the biggest impact, making programmes optimal at higher PPPD (**Figure 2c**). In particular, the Year-round programme goes from never being optimal, to becoming optimal at PPPD of less than £31 assuming on average 360 days of protection from Nirsevimab. Vaccination at 2 months of age reduces the price at which large-scale programmes become optimal from £63 to £13. A lower ICER threshold has a small effect on changing the optimal programme.

### Annual costs of seasonal administration

We calculated the maximum annual cost that one should be willing to pay to ensure that the seasonal programmes remain cost-effective at £20,000/QALY, compared to their year-round counterpart for each PPPD. Assuming a PPPD of between £5-32, then the maximum annual cost that one should be willing to pay to ensure that the Seasonal with catch-up programme remain cost-effective is the threshold value given by the equation: 20887x + 1282463, where x is the PPPD, above which the Year-round programme becomes optimal (Figure 3b). Further, assuming a PPPD between £33–63, then the Year-round programme becomes optimal if the annual cost of seasonal administration exceeds the threshold value given by the equation: 313059x 8710026. Similar formulas exist for the maximum annual costs for x > £63, the threshold value is 524033x 21343865.

**Figure 3.**
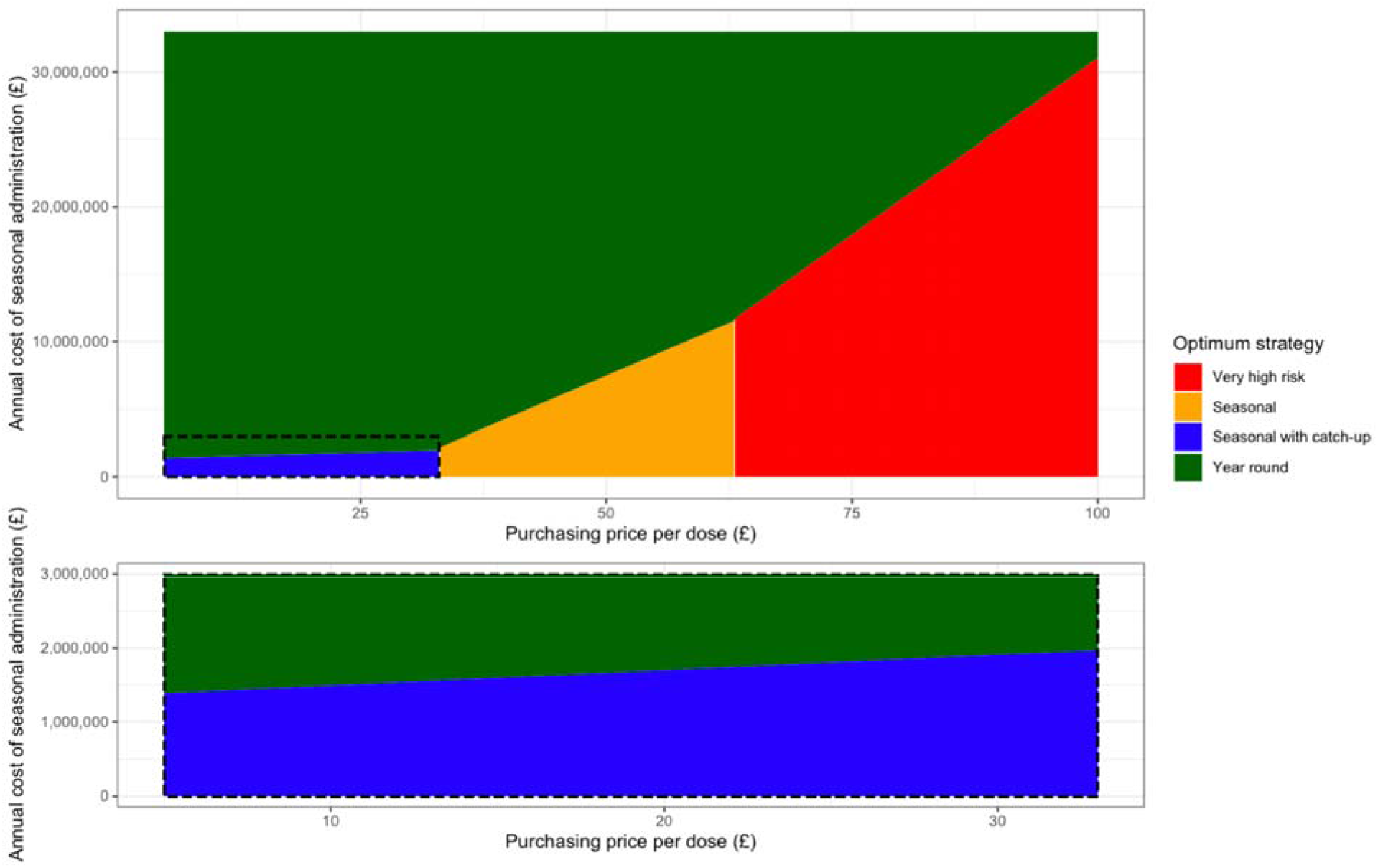
Annual cost of seasonal administration. The impact of the annual cost of seasonal administration and the PPPD on the optimal strategy.

## DISCUSSION

This study used a previously published transmission model to estimate the impact and cost-effectiveness of the monoclonal antibody, Nirsevimab. We evaluated the impact of five different intervention programmes and estimated the optimal strategy given a PPPD. We found that a PPPD of Nirsevimab of £63 or less means that replacing the Palivizumab programme with a Nirsevimab programme targeting all infants seasonally is cost-effective assuming an ICER threshold of 20,000 £/QALY. Specifically, we found between £1–32 PPPD the seasonal with catch-up programme is optimal, and between £33-63 PPPD the seasonal programme is optimal. With our base-case assumptions, the year-round programme and the Seasonal with a NIP-integrated catch-up programme was dominated by the Seasonal with catch-up programme. However, we found that the year-round programme may be cost-effective under two conditions. First, if the duration of protection was 360 days, then the year-round programme was optimal assuming a PPPD of less than £19. Second, if year-round delivery at birth was substantially cheaper than a seasonal programme, then the year-round programme is optimal.

The dynamic transmission model showed that there is little indirect protection from the Nirsevimab programmes. This is because, within the transmission model, the contact matrix used to determine the age-dependent force of infection has very few contacts between infants less than 1 year and other individuals in the population. Consequently, reducing incidence in infants less than one year of age will have a negligible effect on herd protection as they have no significant role in onward transmission. The limited role of infants in the onward transmission of RSV is supported by household surveillance studies of RSV transmission in Kenya, which suggest that older children are responsible for infecting infants within a household.[26] Further, we assumed that the RSV incidence displayed a consistent seasonality as observed prior to 2020. The SARS-CoV-2 pandemic has caused seasonal shifts in RSV seasonality in England and Wales, and it remains unclear if a consistent seasonal peak will return to pre-pandemic level.[18,19] If there is a substantial change in RSV incidence and seasonality over the coming year, then limited inferences can be made from the results of this study on the impact and cost-effectiveness of Nirsevimab.

We derived the immunological assumptions about Nirsevimab from the MELODY trial.[10] However, due to limitations in the study design of this model, it was not possible to parameterise all the information from this trial into the model structure. First, the trial suggests that all-cause medically attended lower respiratory tract infections were reduced by 51% due to Nirsevimab administration, implying a reduction in the incidence of other respiratory diseases than RSV. Though a promising result, a separate, multi-pathogen transmission model would be required to accurately evaluate the impact and cost-effectiveness of this effect. Further, this model does not account for the possibility that RSV may cause longer-term sequelae such as wheezing and asthma in later life, as the evidence for this is not yet completely conclusive [27]. If these sequelae can be prevented by Nirsevimab, then the health benefit and therefore the cost-effectiveness of the Nirsevimab programme will be greater than estimated here. Finally, reduction of RSV cases in infants will reduce RSV-associated antibiotic prescribing. This will decrease the relative costs of these programmes and reduce the negative effects of AMR on other pathogens. Finally, we assume that Nirsevimab protection is lost exponentially, however further studies may show that protection due to Nirsevimab is more sustained compared to an exponential loss assumption. Taken all together, the estimates presented here are likely to underestimate the true impact of Nirsevimab, providing conservative estimates of the cost-effectiveness of these programmes.

This study focuses on evaluating large-scale programmes which target all infants for five different time frames aligned with the manufacturer’s proposed target groups. In this analysis, the VHR programme, i.e. directly replacing the Palivizumab programme with the Nirsevimab, is the optimal programme for a PPPD between £64-£4400. If the PPPD is indeed in this range, then it is unlikely that these large-scale programmes targeting all newborns are cost-effective. However, there is scope to augment the existing Palivizumab-eligible population to include more infants who are high-risk, (e.g. all premature infants, etc.). Under these conditions, calculating the impact of these programmes is important, but to be properly evaluated would require a model stratified by RSV-specific clinical conditions which make up clinically relevant high-risk groups. Consequently, if Nirsevimab is priced higher than expected (i.e. > £100 PPPD), a model with a different structure and intervention strategies may be better suited than the framework presented here.

Although we have considered the difference in logistical challenges between the year-round and seasonal programmes through the annual seasonal implementation costs, there are further logistical differences between different types of seasonal programmes that this analysis has not considered. For example, the seasonal programme (without catch-up) will be logistically easier to implement compared to the seasonal with catch-up programmes. Similarly, between the two catch-up programmes themselves; although the seasonal with catch-up programme dominates the seasonal with a NIP-integrated catch-up programme, the latter is likely to be logistically easier to implement as it is integrated into the existing paediatric immunisation programme and doesn’t require extra recruitment of infants. Economic quantification of these logistical benefits between seasonal programmes is difficult and consequently not possible to parameterise into this disease modelling framework. Once the PPPD of Palivizumab is known however, it may motivate the need for cost estimates for logistical benefits, such that fairer comparisons can be made.

This analysis responds to the JCVI’s need for a cost-effectiveness analysis of Nirsevimab. Our findings suggest that Nirsevimab is likely cost-effective and may be preferable to the existing Palivizumab programme protecting not only those at high-risk but all infants at birth with a large-scale seasonal, or seasonal catch-up programme if the PPPD is £63 or less. If Nirsevimab is commercially licensed, and the PPPD is known, this study provides a guide for decision-makers on the cost-effectiveness of introducing large-scale RSV intervention programmes in infants.

## Data Availability

All data produced are available online at https://github.com/dchodge/nambcea

## Conflicts of Interest

DH, MK, FK, SF, and KA have no conflicts of interest to declare. MJ is an unpaid member of the Respiratory Syncytial Virus Consortium in Europe (RESCEU) and Preparing for RSV Immunisation and Surveillance in Europe (PROMISE). RESCEU and PROMISE have received funding from the Innovative Medicines Initiative 2 Joint Undertaking. This Joint Undertaking receives support from the European Union’s Horizon 2020 research and innovation programme and the European Federation of Pharmaceutical Industries and Associations. Neither MJ nor his research group has received any forms of pecuniary or other support from the pharmaceutical industry.

## Author’s contributions

DH, MJ, and KA were involved in the conceptualisation of the study. DH performed the modelling and analysis, coding, created the figures and wrote the original draft of the manuscript. All authors were involved in the reviewing and editing of the manuscript.

## Role of the funding source

DH was funded by the National Institutes of Health (1R01AI141534-01A1). MK and FK were funded by Innovation Fund of the Joint Federal Committee (grant no. 01VSF18015). FK was funded by Wellcome Trust (grant no. 221303/Z/20/Z). SF was funded by Sir Henry Dale Fellowship jointly funded by the Wellcome Trust and the Royal Society (Grant Number208812/Z/17/Z). MJ was supported by the NIHR HPRU in Modelling and Health Economics (grant code HPRU-2019-NIHR200908) and the NIHR in Immunisation HPRU-2019-NIHR200929). None of the funding sources of any of the authors had involvement in this study. The views expressed are those of the authors and not necessarily those of the UK Department of Health and Social Care, the National Health Service, the National Institute for Health Research (NIHR), or UK Health Security Agency.

## Ethics

No ethical approval was needed

## Data availability

The R code and data needed to replicate this study can be found at https://github.com/dchodge/nambcea.

## Notes

### Competing Interest Statement

The authors have declared no competing interest.

### Funding Statement

This study did not receive any funding

