## Supplementary Material for "Optimal Respiratory Syncytial Virus intervention programmes using Nirsevimab in England and Wales"

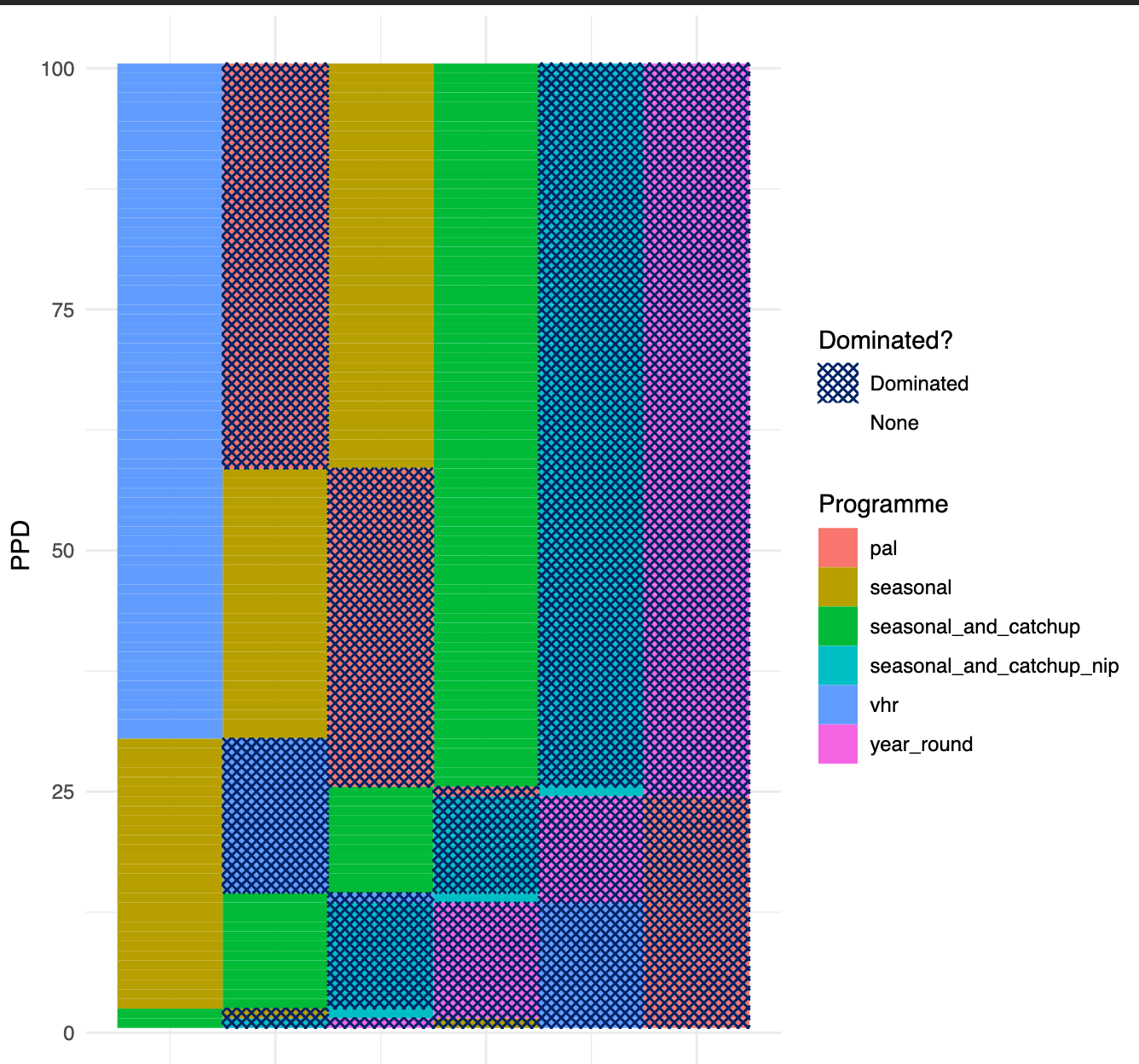

**Figure S1.** Dominance structure for each PPPD. The programmes are ranked in each row in order of cheapest to most expensive (left to right rows) for a given PPPD. Then the dominated programmes (programmes which are more expensive but offer less health benefit than a cheaper programme) on each row are shown through crisscrossed patterns.

| **Parameter** | **Base value** | **Other values** |
| --- | --- | --- |
| Average period of protection (days) | 150 | 250, 360 |
| Coverage | 90% | 70% |
| Age of first vaccination | Birth | 2 months of age |
| ICER threshold (£/QALY) | 20000 | 15000 |

**Table S1**. Parameters associated with the sensitivity
